# A meta-analysis of structural MRI studies of the brain in systemic lupus erythematosus (SLE)

**DOI:** 10.1101/2022.05.13.22275046

**Authors:** Jennifer G. Cox, Marius de Groot, James H. Cole, Steven C. R. Williams, Matthew J. Kempton

## Abstract

**Objective:** A comprehensive search of published literature in brain volumetry was conducted in three autoimmune diseases – Systemic Lupus Erythematosus (SLE), Rheumatoid Arthritis (RA), and Ulcerative Colitis (UC) with the intention of performing a meta-analysis of published data. Due to lack of data in RA and UC the reported meta-analysis was limited to SLE.

**Methods:** The MEDLINE database was searched for studies from 1988 through March 2022. A total of 175 papers met the initial inclusion criteria and 16 were included in a random effects meta-analysis. The reduction in the number of papers included in the final analysis is primarily due to the lack of overlap in measured and reported brain regions.

**Results:** A significantly lower volume was seen in patients with SLE in the hippocampus, corpus callosum and total gray matter volume measurements as compared to age and sex matched controls. There were not enough studies to perform a meta-analysis for RA and UC; instead we include a summary of published volumetric studies.

**Conclusions:** The meta-analyses revealed structural brain abnormalities in patients with SLE suggesting that lower global brain volumes are associated with disease status. This volumetric difference was seen in both the hippocampus and corpus callosum and total gray matter volume measurements. These results indicate both gray and white matter involvement in SLE and suggest there may be both localised and global reductions in brain volume.

**KEY MESSAGES:** *What’s already known on this topic:*

- Central nervous system effects of lupus are common, however, agreement on principally affected neuroanatomical regions is lacking.

*What this study adds:*

- This study combines the volumetric neuroimaging data from previously published SLE literature in a meta-analysis. The hippocampus, corpus callosum and total gray matter volume in patients with SLE is smaller than in age and sex matched controls. Additionally, a summary of published data in RA and UC is provided.

*How this study might affect research, practice or policy:*

- This is the first meta-analysis on neuroimaging studies of volume differences in SLE. The regions identified can inform further research on disease progression and therapy evaluation targeted at brain volumetric changes in SLE.
- These results provide specific regions of interest to further explore in the central treatment and management of SLE. While these regions are shown to be directly affected, additional brain regions may be implicated. Further research to understand the potential link between these volumetric measurements and behavioural/cognitive changes observed in patients with lupus is warranted.

## INTRODUCTION

The term autoimmune disease encompasses a large and heterogeneous group of disorders that afflict specific target tissues. The distinction between these different diseases can be minimal with significant overlap between various autoimmune diseases. Additionally, the presence of multiple autoimmune disease diagnoses, or polyautoimmunity, both in individuals and within families is well documented and can lead to overlap in symptom presentation. [1]

Traditionally, both in clinical drug development and clinical practice, the primary focus has been in the peripheral management of disease burden. However, many patients report high levels of central nervous system symptoms such as pain, fatigue, and depression. [2, 3] These reports of central manifestations have contributed to the increased interest in further understanding the neuro-immune axis. Autoimmune diseases represent an interesting opportunity to explore whether systemic inflammation from these autoimmune pathways have a central effect and, if so, whether there is a common or discrete central effect caused by different autoimmune diseases.

Individual neuroimaging studies have reported associations with autoimmune disorders, however, there have been inconsistent findings. Here, we sought to meta-analyse the existing reports of brain volumetry in three different autoimmune diseases, Systemic Lupus Erythematosus (SLE), Rheumatoid Arthritis (RA) and Ulcerative Colitis (UC) and to summarise the results of various reported brain regions.

## METHOD

### Database of Imaging Studies in SLE, RA and UC

A MEDLINE search of studies published from 1988 through March 2022 was performed, combining Medical Subject Heading (MeSH) terms and free text searches. The search was completed in March 2022.

Prior to any data extraction this study was registered on the PROSPERO website under the registration ID:CRD42021210020. [4] Full search terms can be found on PROSPERO and in the supplementary materials. Volumetric data was then extracted for all reported regions for all three diseases. A meta-analysis was then conducted, reported following the PRISMA checklist. [5, 6]

Published studies that measured brain volumetry using MRI in patients with SLE, RA and UC which had a healthy control group were included in the database. To be included in this meta-analysis papers had to be in English and have reported quantitative structural neuroimaging results. All case studies/case series were excluded.

A total of 1070 publications were identified, of which 38 met final inclusion criteria. After data extraction this was reduced to 16 papers. This is primarily due to the lack of overlap in reported regions and insufficient volume of published data available in UC and RA. Further information on the UC and RA search results, including a summary of published volumetric literature can be found in the results section.

Further details regarding the study identification process are provide in the PRISMA inclusion flow chart below (Figure 1).

**Figure 1:**
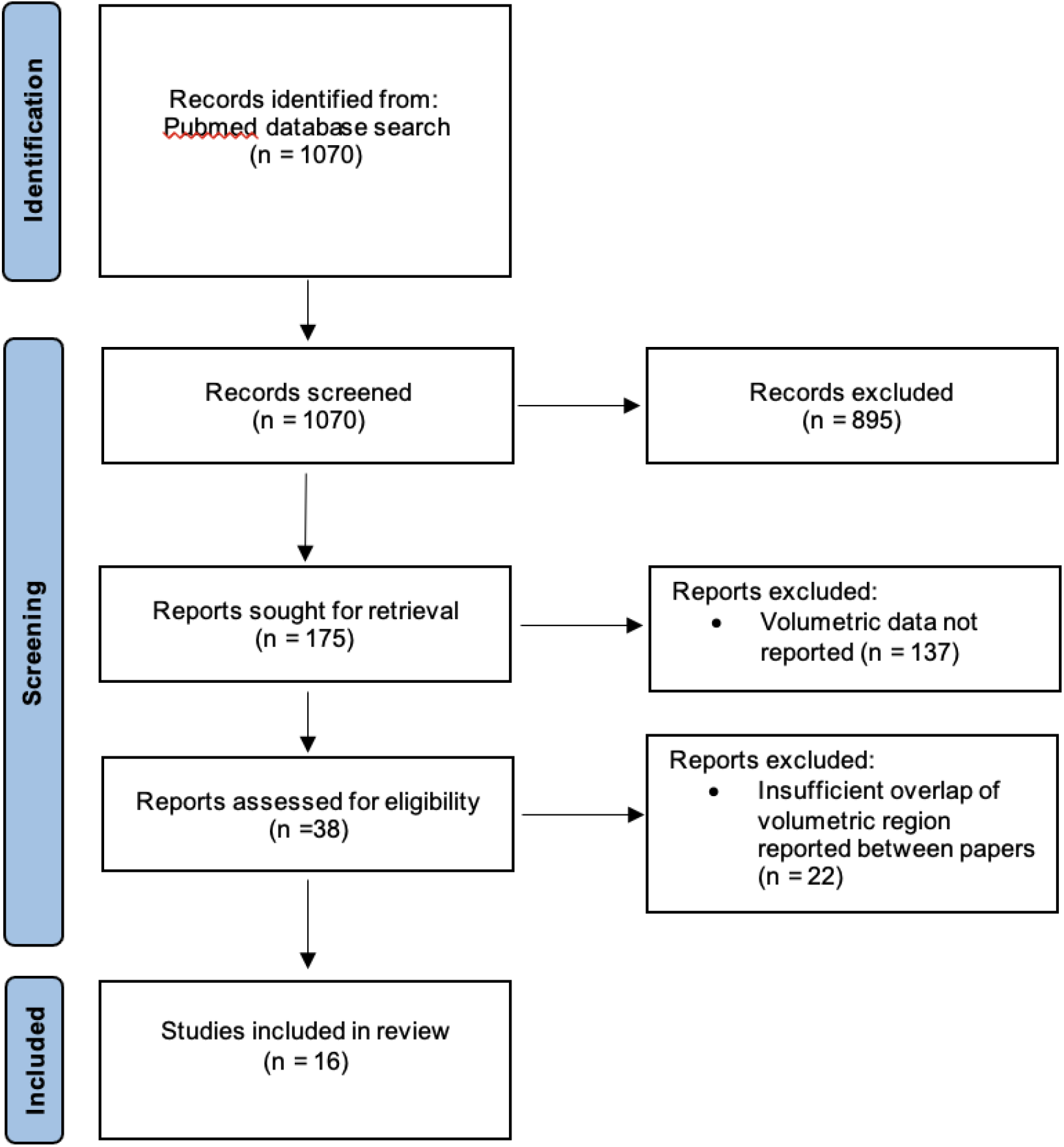
Adapted PRISMA Flowchart Showing Study Inclusion

Patients and the public were not involved in the design, conduct, reporting or dissemination plans of the present research.

### Autoimmune Disease Volumetry Meta-Analysis

From the 6 UC papers and 6 RA papers reporting volumetric neuroimaging data there was not sufficient overlap between the reported regions to power a meta-analysis. We therefore decided to exclude these disorders from the meta-analysis and focus on SLE, however we have included a review of these UC and RA studies in the results section.

One study was excluded as the results of the statistical test did not match the reported mean and standard deviation. This led to exclusion of whole brain volume in NPSLE from this meta-analysis after the data extraction phase as there were then less than 3 studies reporting that region of interest.

The list of the 16 studies included in this meta-analysis can be found in Table 1. Of these, we selected the 5 regions that were reported by three or more studies to ensure that each meta-analysis was sufficiently powered.

**Table 1:** Studies included in the SLE Meta-Analysis

| Study | No. of SLE Patients | No. of Controls | Brain Regions Reported |
| --- | --- | --- | --- |
| Kozora et al, <sup>[21]</sup> 2005 | 7 | 7 | Hippocampus |
| Appenzeller et al, <sup>[22]</sup> 2006 | 107 | 40 | Hippocampus |
| Shapira-Lichter et al, <sup>[23]</sup> 2013 | 12 | 11 | Hippocampus |
| Lapa et al, <sup>[24]</sup> 2017 | 54 | 56 | Hippocampus |
| Cannerfelt et al, <sup>[25]</sup> 2018 | 27 | 25 | Hippocampus, Corpus Callosum |
| Liu et al, <sup>[26]</sup> 2020 | 85 | 77 | Hippocampus |
| Kamintsky et al, <sup>[38]</sup> 2020 | 65 | 9 | Hippocampus |
| Appenzeller et al, <sup>[27]</sup> 2005 | 115 | 44 | Corpus Callosum |
| Lee et al, <sup>[28]</sup> 2015 | 12 | 22 | Corpus Callosum |
| Tamires Lapa et al, <sup>[29]</sup> 2016 | 76 | 66 | Corpus Callosum |
| Emmer et al, <sup>[30]</sup> 2006 | 21 | 12 | Total Gray Matter |
| Cagnoli et al, <sup>[31]</sup> 2012 | 18 | 18 | Total Gray Matter |
| Tjensvoll et al, <sup>[32]</sup> 2016 | 53 | 53 | Total Gray Matter |
| Liu et al, <sup>[33]</sup> 2018 | 89 | 84 | Total Gray Matter |
| Zivadinov et al, <sup>[34]</sup> 2013 | 26 | 36 | Total Gray Matter |
| Cesar et al, <sup>[35]</sup> 2015 | 23 | 43 | Total Gray Matter |

The majority of studies report absolute volume measures; however, one study reported hippocampal volume as a percentage of intracranial volume (ICV). All measurements have been included in the meta-analysis; however, because combining measures may increase heterogeneity, an additional analysis was carried out with volume measures only. The results of this additional analysis are included in the hippocampal volume section of the results.

We identified 4 papers reporting corpus callosum volume, however, one paper reported the median and range rather than the mean and standard deviation. To allow the inclusion of this study we estimated the standard deviation and mean based on the reported median, range and sample size; the calculation was based on an established methodology for meta-analyses.[15]

To calculate an effect size from each study we used Hedges’ g, which is the Cohen’s effect size with a correction for bias from small samples. [16] Outcome measures were combined using a random-effects inverse-weighted variance model. [17] The Cochran Q statistic was calculated to examine the heterogeneity between studies. [18] The I^2^ statistic was also calculated, which is equal to the percentage of total variation between studies due to heterogeneity. [19] The effect of small-study bias (which may include publication bias) was investigated, however, given the small number of studies meta-analysed for each region this test was under powered and therefore not reported.

### SLE Patient population

Given the observed manifestation of a wide range of neuropsychiatric symptoms, a standardised nomenclature system for neuropsychiatric syndromes of systemic lupus erythematosus (NPSLE) has been created by the American College of Rheumatology. [20] Since the initial publication of these criteria there have been other criteria proposed and in practice the categorisation of NPSLE varies greatly between researchers and publications. For the purposes of this meta-analysis, patient groups designated as NPSLE or as having neuropsychiatric manifestations have been analysed separately to patients labelled as SLE. This is particularly important as some of the included and related literature recruited an SLE, an NPSLE and a control group. All papers included in this meta-analysis were published after the publication of this initial NPSLE criteria in 1999.

## RESULTS

Four regions were reported by a sufficient number of studies to be included in this meta-analysis. These were the hippocampus, corpus callosum and total gray matter volume in the SLE population and total gray matter volume in the NPSLE population. The 16 studies included in the meta-analysis and associated brain regions are listed in Table 1.

### Meta-analysis of Hippocampal Volume in SLE

Of the 4 regions identified in this meta-analysis, hippocampal volume was reported the most frequently. In all papers, right and left hippocampal volume was reported and was meta-analysed separately. In reviewing the meta-analysis results the Lapa et al, 2017 paper appeared as an outlier (effect size of outlier, -4.3; effect size range of the remaining 5 studies, -1.21 to 0.24). We present results excluding and including the outlier for both the left (Figure 2) and right (Figure 3) hippocampal volumes. Both with and without the outlier, pooled effects sizes show significantly smaller right and left hippocampal volume in SLE patients compared to controls. Results for all brain regions including the hippocampus are reported in Table 2.

**Table 2:** Meta-analysis of Continuous Data Comparing Patients with SLE vs Controls

| Region | Studies (N) | SLE or NPSLE Patients vs Control Subjects (N/N) | Effect Size |  |  | Heterogeneity<sup>a</sup> |  |
| --- | --- | --- | --- | --- | --- | --- | --- |
|  |  |  | Hedges g | 95% CI | p-value | I<sup>2</sup> % | p-value |
| **Hippocampus (right)** | 7 | 357/225 | -0.98 | -1.89, -0.06 | 0.037 | 94.8 | <0.001 |
| **Hippocampus (left)** | 7 | 357/225 | -1.06 | -1.98, -0.13 | 0.025 | 94.8 | <0.001 |
| **Hippocampus (right) – outlier excluded** | 6 | 303/169 | -0.49 | -0.88, -0.11 | 0.013 | 65.5 | 0.013 |
| **Hippocampus (left) – outlier excluded** | 6 | 303/169 | -0.56 | -0.99, -0.14 | 0.009 | 70.9 | 0.004 |
| **Corpus Callosum** | 4 | 230/157 | -0.56 | -1.11, -0.01 | 0.045 | 82.2 | 0.001 |
| **Total GM Volume (SLE)** | 4 | 181/167 | -0.46 | -0.84, -0.08 | 0.018 | 60.5 | 0.055 |
| Total GM Volume (NPSLE) | 4 | 106/109 | -0.28 | -0.87, 0.30 | 0.343 | 75.3 | 0.007 |

<sup>a</sup> Publication bias was not reported as the number of studies was not large enough to sufficiently power this calculation.
Regions with significant findings shown in bold

**Figure 2:**
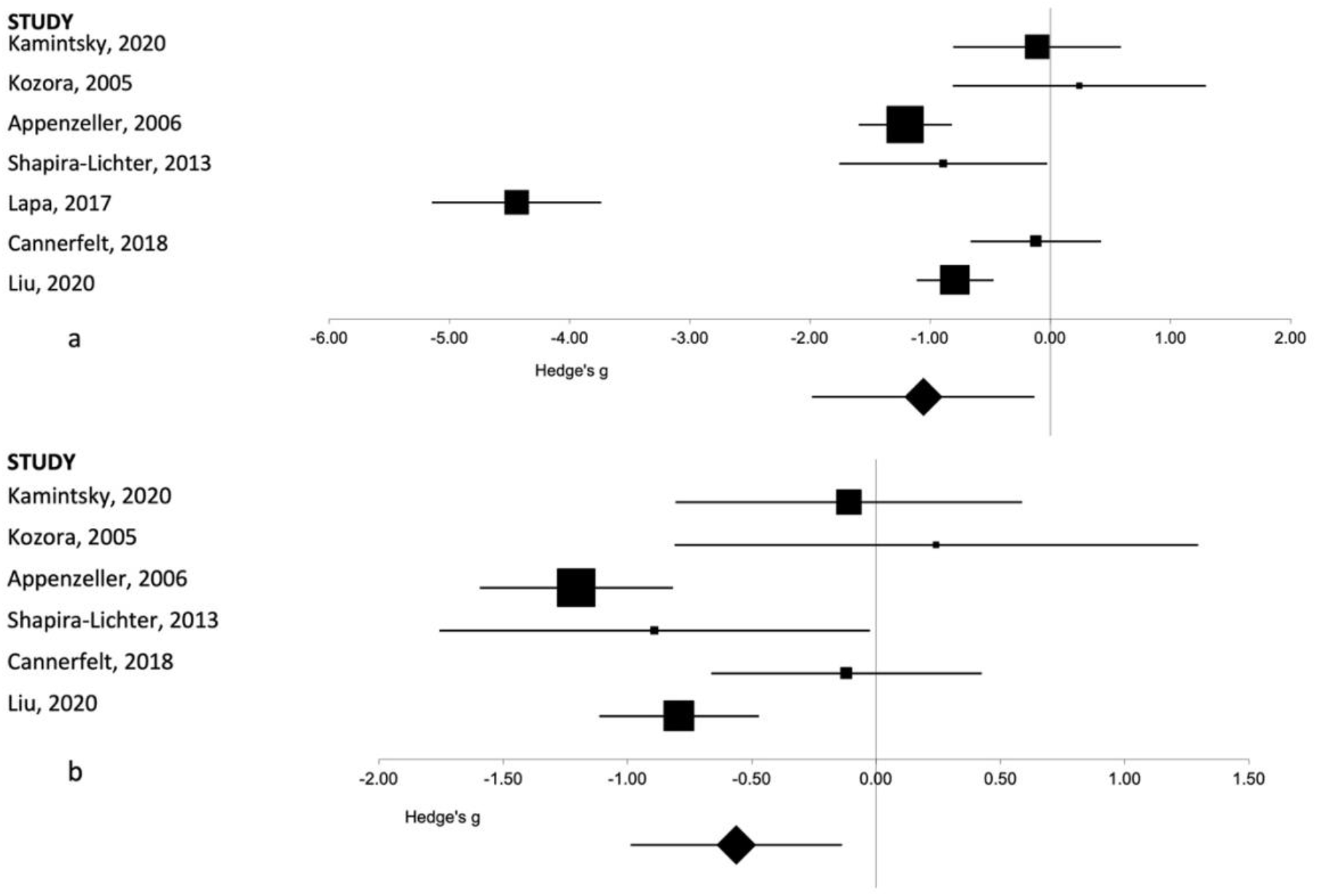
Forest plot representing the meta-analysis of the left hippocampal volume in patients with SLE as compared to healthy controls. The size of the squares in the plot represents the weight of each study in the analysis, the error bars correspond to the 95% confidence intervals, the diamond shape represents the pooled effect size. (a) Results with the outlier included. (b) Results with the outlier excluded.

**Figure 3:**
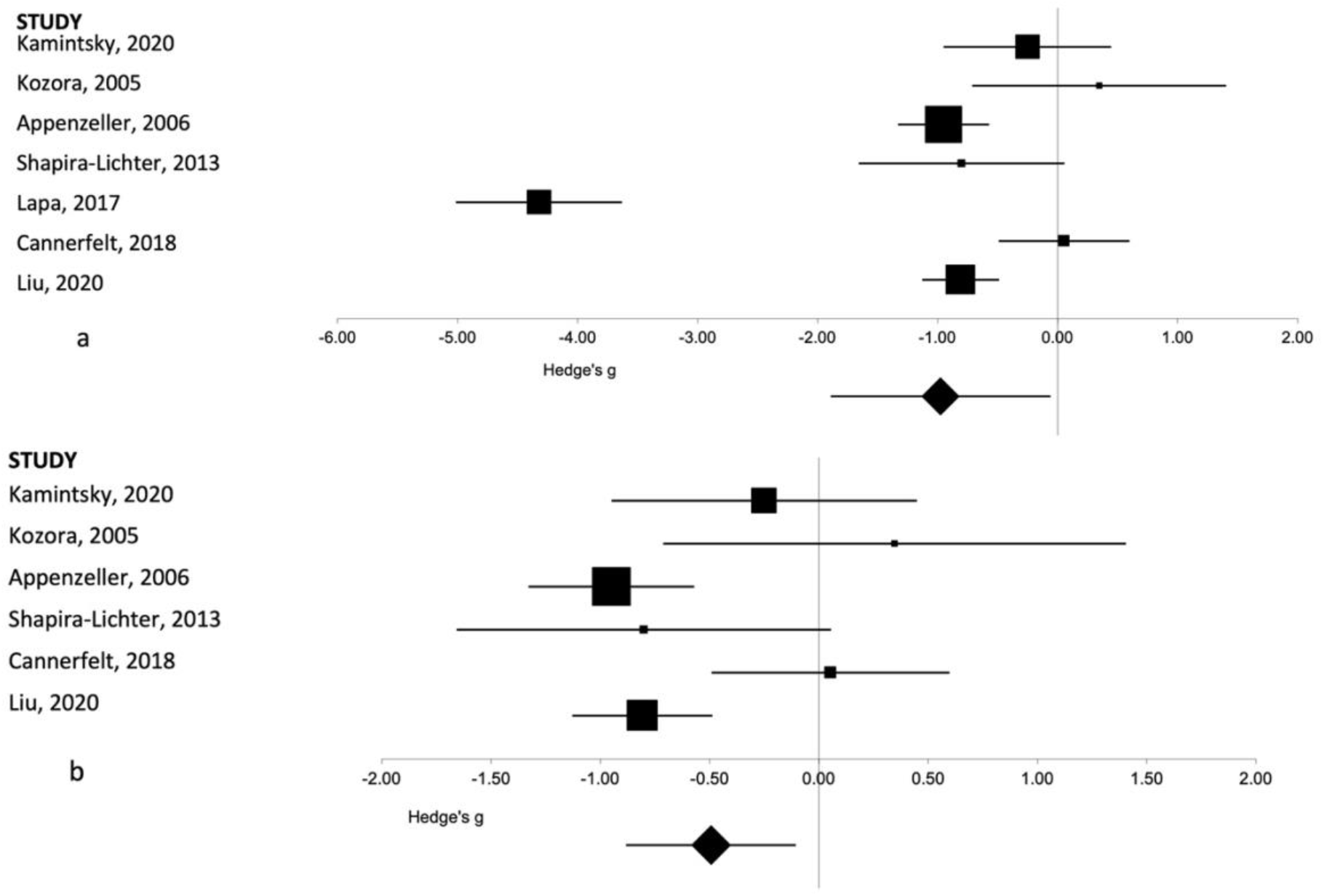
Forest plot representing the meta-analysis of the right hippocampal volume in patients with SLE as compared to healthy controls. The size of the squares in the plot represents the weight of each study in the analysis, the error bars correspond to the 95% confidence intervals, the diamond shape represents the pooled effect size. (a) Results with the outlier included. (b) Results with the outlier excluded.

The results from the Kamintsky et al, 2020 paper were reported as a percentage of ICV rather than an absolute volume measurement. An additional analysis was performed excluding this paper, however, this did not materially affect the overall results. For the left hippocampus this resulted in an overall effect size of -1.21 (95%CI -2.25 to -0.18, p=0.02) rather than -1.06 (95%CI -1.98 to -0.13, p=0.03) seen with this data included. For the right hippocampal volume this results in an overall effect size of -1.1 (95%CI -2.13 to -0.06, p=0.04) versus - 0.98 (95%CI -1.89 to -0.06, p=0.04).

### Meta-analysis of Corpus Callosum Volume in SLE

As described in the methods section, Tamires Lapa et al, ^[29]^ 2016 reported median and range therefore an estimated mean was used for those values in this meta-analysis. The resulting estimated mean values were very close to the reported medians (median SLE, 11.6 cm^3^; estimated mean SLE, 11.6 cm^3^; median controls, 13.7 cm^3^; estimated mean controls, 13.9 cm^3^) suggesting a symmetric and possibly normal distribution of the data and providing confidence in the estimated values. Overall, a lower whole corpus callosum volume was seen in patients with SLE as compared to controls as seen in the overall pooled effect size. These results are shown in Figure 4. Full results are reported in Table 2.

**Figure 4:**
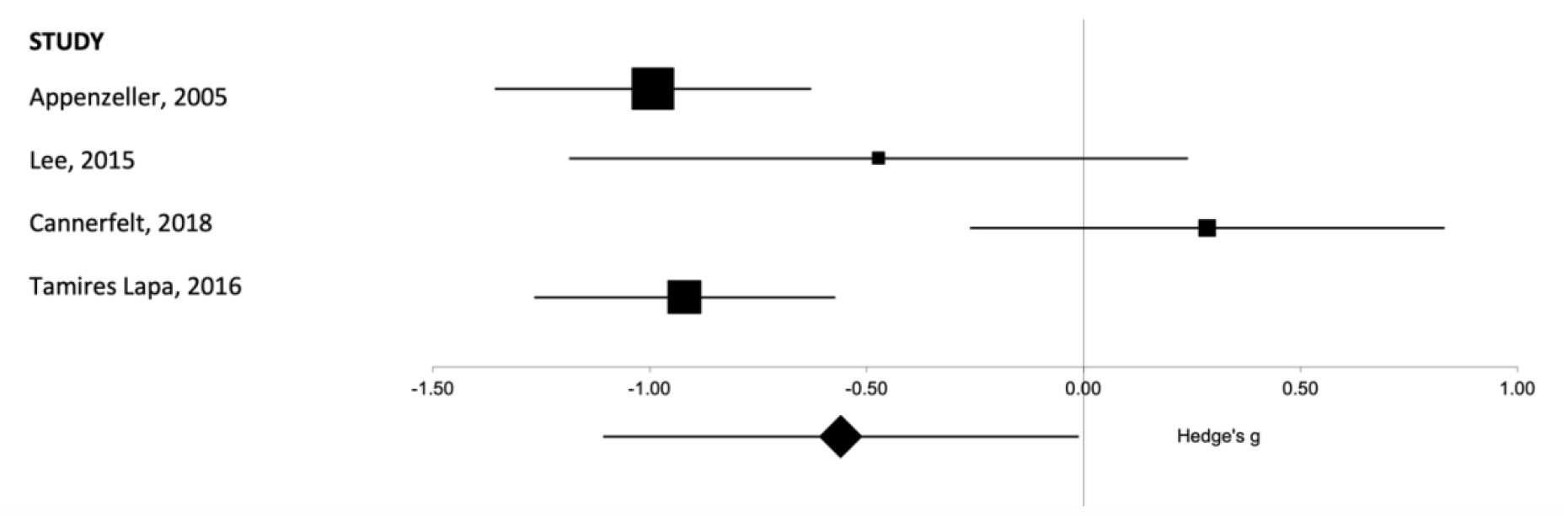
Forest plot representing the meta-analysis of the corpus callosum volume in patients with SLE as compared to healthy controls. The size of the squares in the plot represents the weight of each study in the analysis, error bars correspond to the 95% confidence intervals, the diamond shape represents the pooled effect size.

### Meta-analysis of Total Gray Matter Volume in SLE and NPSLE

For total gray matter volume there were 6 studies in total and enough reported data to separately analyse SLE versus controls, and NPSLE versus control values. Two of the papers had an NPSLE, SLE and a healthy control arm. There were 2 additional papers that reported just SLE versus healthy controls, and 2 additional papers that reported NPSLE versus healthy controls. This allowed 4 studies to be included in a SLE vs healthy control meta-analysis and 4 studies to be included in the NPSLE vs healthy control meta-analysis. There was a significantly lower gray matter volume seen in SLE patients as compared to controls (figure 5a), however, this same result was not seen in NPSLE subjects compared to control subjects (figure 5b). Full results are provided in Table 2.

**Figure 5:**
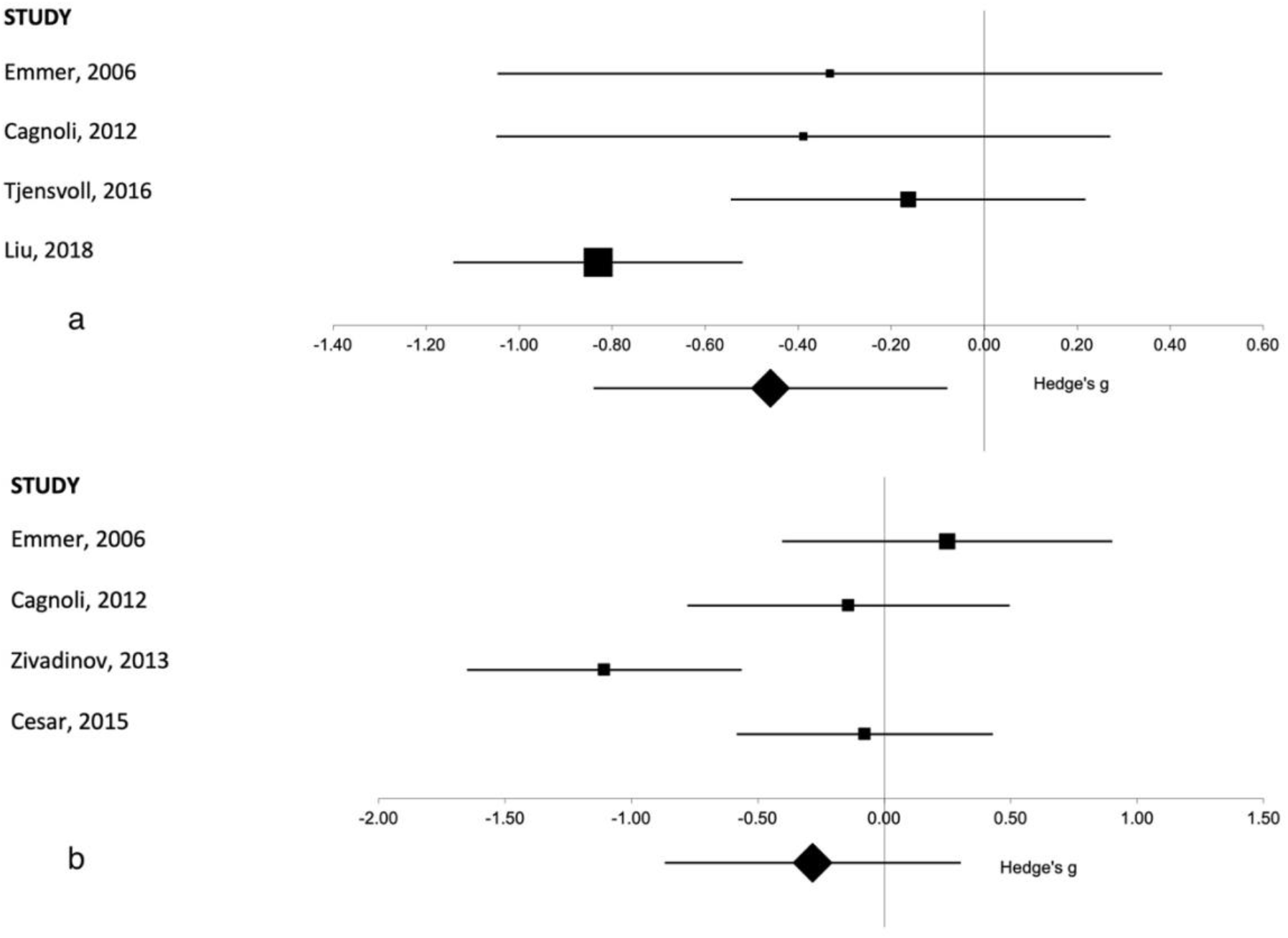
Forest plot representing the meta-analysis of the total gray matter volume in patients (a) with SLE as compared to healthy controls and in patients (b) with NPSLE as compared to healthy controls. The size of the squares in the plot represents the weight of each study in the analysis, error bars correspond to the 95% confidence intervals, the diamond shape represents the pooled effect size.

### Summary of published RA and UC data

While there were insufficient volumetric data available to perform a meta-analysis in RA and UC, there were multiple published papers examining central effects in these diseases.

In RA, there was a confirmed recognition of a central effect on brain volumes, with different brain regions reported between different studies. Two separate research groups reported associations between disease activity or disease duration with lower gray matter volume. [7,8] Schrepf and colleagues utilised inflammatory markers such as Erythrocyte Sedimentation Rate (ESR) and C-reactive protein (CRP) and linked increased peripheral inflammation to lower GM volumes particularly in the cerebellum. [7] Bekkelund and colleagues noted a larger ventricle-to-brain ratio and smaller midsagittal cerebellar areas correlated with longer disease duration. [8] Other studies reported lower olfactory bulb volumes in RA patients and differences in the subcortical gray matter potentially reflecting generalised volume reductions or differences in brain development in patients with RA. [9,10] One report found greater putamen gray matter volumes correlated with higher fatigue scores in RA patients; the authors speculated this being a potentially compensatory mechanism due to low putamen activity. [11]

There were fewer published studies in UC. This may be due to inconsistent use of terminology. For example, some studies report results from patients with inflammatory bowel disease (IBD), an umbrella term most often including both Crohn’s Disease (CD) and UC. Two reports from Agostini and colleagues examined brain morphological abnormalities in different patient populations within IBD. Despite comparable sample sizes, results indicated a significantly lower GM volume in a CD patient group, however, they did not find similar results in a UC group. Agostini et al state that this could have potentially been a function of the overall mild clinical course of disease and remission status of the recruited patient sample. [12, 13] Similarly, Zikou and colleagues reported lower GM volumes specifically in the fusiform gyrus, the right middle frontal gyrus and the left superior parietal gryus in an IBD patient group. They did not specify what percentage of their patient group had UC. [14] The most recent paper published in 2021 by Zhang and colleagues, reports the most extensive list of volumetric regions measured in a UC specific population. They report areas of both increased and decreased gray matter volume associated with both presence of the disease as compared to healthy controls, and with disease remission status. [37] Further volumetric studies need to be conducted in a UC specific population to understand whether there are consistent volumetric differences present within this population.

## Discussion

In patients with SLE compared with controls, we found lower volume of the right and left hippocampus, corpus callosum cross-sectional area, and total gray matter. These volumetric differences are consistent with previously reported findings from individual studies and indicate both white matter and gray matter involvement. [26, 29, 33] When comparing patients with NPSLE to controls there was no significant difference in total gray matter volume.

Heterogeneity was generally high for brain volume regions, particularly for the hippocampus although this was reduced when we excluded an outlier. It is not immediately clear why this outlier is so significantly different from the other reported studies. We speculate that this is due to differences in image acquisition. Four of the other five studies reported used a 3D sequence specifically for their T1-weighted volumetric analysis.

The NPSLE finding in gray matter is surprising given the difference observed in the pooled SLE data as compared to control subjects, particularly given the overlap of studies, with half of the studies used to pool data having a separate NPSLE and SLE arm. However, it is important to note that this finding was from a relatively small sample size, both in the number of reported studies and in the overall sample size of the pooled data, and therefore requires further investigation. Our meta-analysis did not compare SLE versus NPSLE patients as there were not enough studies making direct comparisons. In the two studies that did recruit separate arms there were no significant differences observed between SLE and NPSLE patients.

As hypothesized, systemic inflammation in SLE is associated with lower brain volumes. The relationship between volume differences of specific brain regions and reported cognitive findings and neuropsychiatric manifestations is still not well understood. There is some indication that volume reductions may precede cognitive impairment in SLE and therefore could be a predictive marker for future central effects. However, this would require a prospective, longitudinal study to further investigate if this is potentially causal relationship and further contextualise the predictive value of this measure both in patients with and without neuropsychiatric involvement.

One limitation to the current meta-analysis was the low number of individual studies included and hence the relatively small size of the overall pooled sample. While there was enough commonality in reported brain regions between papers to perform this meta-analysis there were a number of regions that had been analysed and published, though with insufficient overlap between studies to meta-analyse. This includes many additional subcortical regions such as the thalamus, putamen and amygdala. There could also potentially be alternative brain regions that are impacted by SLE that are therefore not captured here. Additionally, given the whole gray matter volume effect seen in SLE patients, it is unclear whether these effects are localised to specific regions or part of a larger, more widespread pattern of brain atrophy.

An additional limitation when interpreting these data is the known effect of corticosteroids on neurostructural volumes, particularly the hippocampus.[36] Medication use was not taken into account in this meta-analysis specifically, however, it was discussed and incorporated into individual studies included in this analysis. We attempted to document this and include this as a sub-analysis, but corticosteroid usage was not consistently reported across all studies. Corticosteroids are a first line therapy in treating various forms of lupus and therefore the observed difference in volume in SLE patients cannot necessarily be attributed solely to disease status alone. Further research is necessary with a larger sample size, ideally with a control arm including people who regularly receive corticosteroids but do not have an autoimmune diagnosis (e.g., asthma patients).

To our knowledge, this is the first meta-analysis to look at volumetric neurostructural changes in SLE and provides a strong basis for further research both in the regions reported here and potentially looking at additional areas of the brain in future. The existence of some peer reviewed published data, but lack of volume of data for the purposes of meta-analysis in UC and RA indicates community interest in further understanding the central effect of these diseases. Future studies comparing these populations to controls and to each other could provide valuable information about UC and RA individually and about autoimmune diseases more generally.

## Supporting information

Supplementary Search Strategy Document

## Data Availability

All data produced in the present work are contained in the manuscript

## Acknowledgementsand Affiliations

From the Department of Neuroimaging and the Department of Psychosis Studies, Institute of Psychiatry, Psychology, and Neuroscience, King’s College London

Address correspondence to Jennifer Cox.

Jennifer Cox is an industry funded PhD student funded by GlaxoSmithKline.

Dr. de Groot is an employee of, and holds shares in GlaxoSmithKline (GSK). GSK had no role in the design or conduct of the study.

Dr. Kempton was funded by an MRC Career Development Fellowship (grant MR/J008915/1). The authors acknowledge financial support from the Wellcome Trust and the Engineering and Physical Sciences Research Council for the King’s Medical Engineering Centre and the National Institute for Health Research (NIHR) Biomedical Research Centre at South London and Maudsley NHS Foundation Trust and King’s College London.

The views expressed here are those of the authors and not necessarily those of the NHS, the NIHR, or the Department of Health.

Dr. Williams has received research funding from Bionomics, Eli Lilly, the Engineering and Physical Sciences Research Council, GlaxoSmithKline, Johnson & Johnson, Lundbeck, the National Institute of Health Research, Pfizer, Takeda, and Wellcome Trust. The other authors report no financial relationships with commercial interests.

