## Supplementary Search Strategy Document for "A meta-analysis of structural MRI studies of the brain in systemic lupus erythematosus (SLE)"

### Search Strategy for Literature Review and Meta-Analysis:

#### Search Terms:

((Systemic Lupus Erythematosus) OR (SLE) OR (lupus) OR (neuropsychiatric systemic lupus erythematosus) OR (NPSLE) OR (lupus vasculitis) OR (rheumatoid arthritis) OR (ulcerative colitis)) AND (volume OR Hippocampus OR sulcus OR gyrus OR putamen OR caudate OR ventricle OR (prefrontal) OR parietal OR occipital OR cingulate OR amygdala OR ("white matter") OR ("grey matter") OR ("gray matter") OR cortex) AND MRI

#### Database Used:

PubMed

#### Original Search Date:

18 November 2020

#### Updated Search Date:

03 March 2022

#### Restrictions:

- English language only
- Must include subjects with Systemic Lupus Erythematosus (SLE), Rheumatoid Arthritis (RA) or Ulcerative Colitis (UC)
- Have reported structural neuroimaging measures available
- Include a control arm in the study design
- All case studies/case series excluded
